# MRI based Kidney Radiomics analysis during chronic lithium treatment: validation of a texture index associated with decreased kidney function

**DOI:** 10.1101/2021.10.23.21265420

**Authors:** Paul Beunon, Maxime Barat, Anthony Dohan, Lynda Cheddani, Lisa Males, Pedro Fernandez, Bruno Etain, Frank Bellivier, François Vrtovsnik, Emmanuelle Vidal-Petiot, Antoine Khalil, Martin Flamant, Nahid Tabibzadeh

## Abstract

**Background:** Chronic lithium therapy is associated with an increased risk of chronic kidney disease (CKD). Lithium nephrotoxicity is slowly progressive and difficult to detect at early stages. The aim of this study was to identify specific image texture changes of kidneys as possible imaging biomarkers of decreased measured glomerular filtration rate (mGFR) using radiomic analysis of T2 weighted imaging magnetic resonance imaging.

**Methods:** One hundred and nine patients treated with lithium carbonate were evaluated including mGFR and Kidney MRI, with T2 weighted sequence single-shot fast spin-echo. Computed radiomic analysis was performed after a manual kidney segmentation. Significant features were selected to build a radiomic signature using multivariable Cox analysis to detect a decreased mGFR <60 ml/min/1.73m^2^. The texture index was internally validated using a training and a validation cohort.

**Results:** Texture analysis index was able to detect a decrease in mGFR, with an Area Under the Curve (AUC) of 0.85 in the training cohort and 0.71 in the validation cohort. Patients with a texture index below the median were older (59 [42-66] versus 46 [34-54] years, p=0.001), with longer treatment duration (10 [3-22] versus 6 [2-10] years, p=0.02), and a lower mGFR (66 [46-84] versus 83 [71-94] ml/min/1.73m^2^, p<0.001). Texture analysis index was independently and negatively associated with age (β= -0.004 ±0.001, p<0.001), serum vasopressin (−0.005 ± 0.002, p=0.02), lithium treatment duration (−0.01 ± 0.003, p=0.001), with a significant interaction between lithium treatment duration and mGFR (p=0.02).

**Conclusion:** A renal texture index was developed and validated among patients treated with lithium carbonate associated with a decreased mGFR. This index might be relevant in the diagnosis and prognosis of lithium-induced renal toxicity.

## Introduction

Lithium carbonate is the cornerstone long-term treatment of bipolar disorder. The efficacy of lithium is proven in the prevention and treatment of acute maniac and depressive episodes as well as in the prevention of suicidal risk (1–3). However, this efficacy is counterbalanced by potentially serious adverse events. Even at therapeutic ranges, long term lithium treatment might lead to polyuria and polydipsia, related to impaired urine concentrating ability, and eventually to an increased risk of chronic kidney disease (CKD) (4–6).

Specific causal attribution of CKD to lithium treatment in this population is still controversial. Although some authors have related CKD to comorbidities such as ageing, hypertension and metabolic syndrome in these patients (7), late-stage lithium-associated CKD is characterized by a typical pattern associating tubular atrophy, interstitial fibrosis and diffuse cortical and/or medullary microcysts than can be evidenced on renal magnetic resonance imaging (MRI) (8–11). Moreover, experimental models have shown structural changes early after lithium initiation, characterized by cellular proliferation within renal collecting ducts (12,13). To date, these early changes cannot be detected through non-invasive tests. Yet, the detection of lithium-induced early nephrotoxicity is of major importance when facing the issue of lithium discontinuation and would be help weighing the risk/benefit ratio.

Radiomics is a growing field of research consisting in the mathematical integration of high throughput data produced by medical imaging (14). The image texture is analyzed at the pixel scale, allowing the computation of multiple data. Radiomic analyses have been shown to accurately detect the degree of cancer cell proliferation and heterogeneity within the tissue, hence determining the prognosis of solid tumors (14). It might thus represent a noninvasive surrogate biomarker of histological findings, potentially allowing personalized management of patients.

In this view, we hypothesized that non-invasive texture-based imaging biomarkers could provide quantifiable specific modification in patients treated with lithium carbonate and that it could detect a early decrease in measured glomerular filtration rate (GFR) in this population.

We thus aimed at analyzing kidneys of patients treated with lithium using MRI radiomics to internally validate the technique and evaluating the correlates of kidney texture in this population.

## Methods

### Design

One hundred and fourteen patients on chronic lithium treatment (at least 6 months) with available kidney MRI with T2-weighed coronal sequences were retrospectively included in this monocentric study. Five patients were excluded from the analysis due to artifacts (n=2), inappropriate sequences (n=1), incomplete test (n=1), and solitary kidney (n=1) (study flowchart, figure 1).

**Figure 1.**
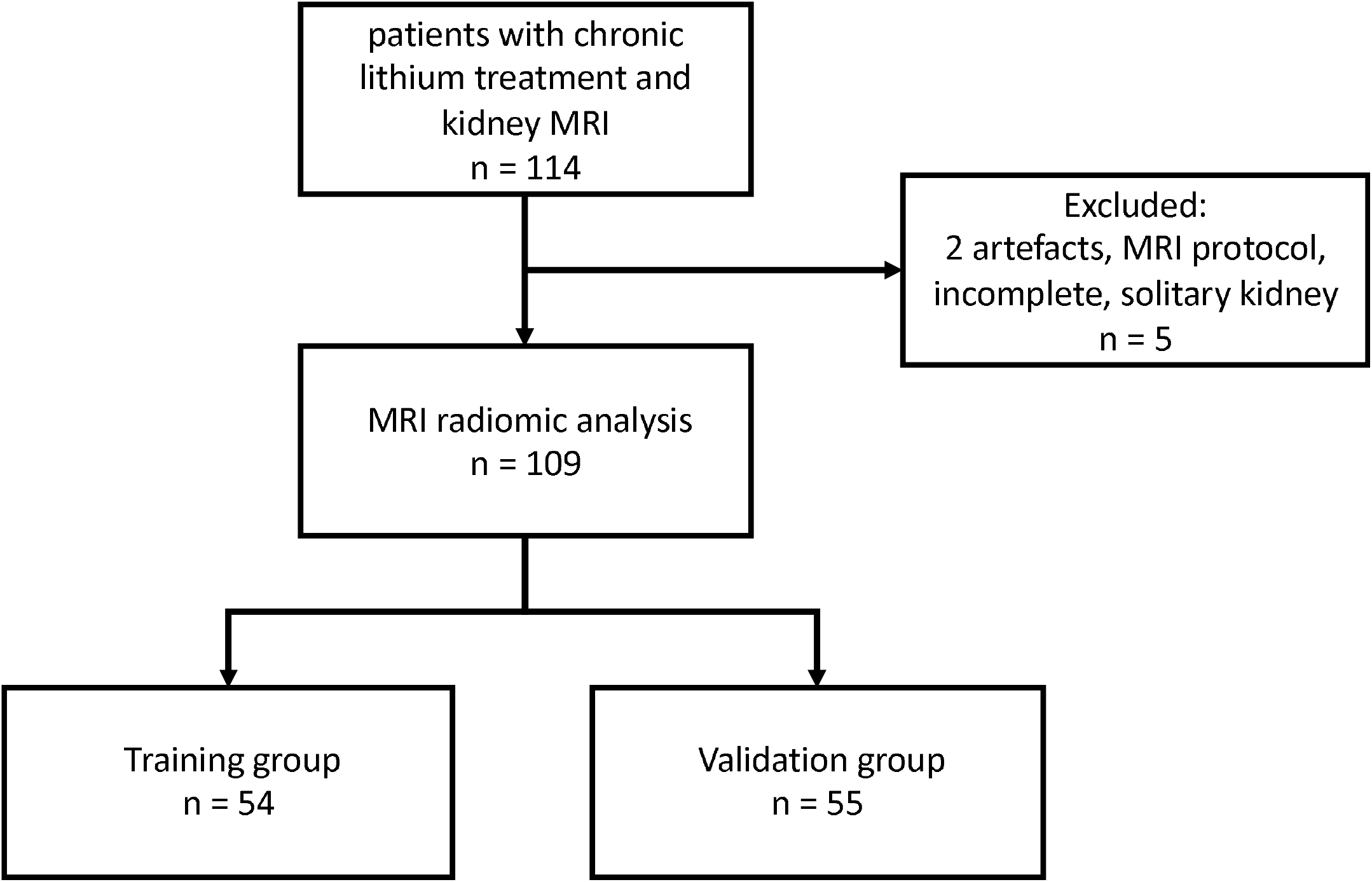
Study Flowchart.

### Population

From March 2015 to December 2020, 109 adult patients were referred by psychiatrists to the Department of Renal Physiology for systematic check-up with nephrologists. Eligible patients were ≥ 18 years of age at inclusion, with various durations of lithium treatment and no dialysis or kidney transplantation history.

All patients provided written informed consent before inclusion in the study cohort. The study was approved by the local ethics committee (Institutional Review Board CER-2021-74) and Helsinki’s Declaration ethical statements were respected.

### Data collection and measurements

During a half-day in-person visit, clinical and biological parameters were collected. GFR was measured (mGFR) by urinary clearance of 99Tc-DTPA or 51Cr-EDTA (Curium, Saclay, France and GE Healthcare, Velizy, France respectively) as previously described (15). Plasma lithium concentration was the last available measurement during the last 6 months. Serum vasopressin was measured in the Renal Physiology Laboratory of Hôpital Tenon, France, with a standardized and validated protocol as described by Ho *et al* (16). Desmopressin (dDAVP) was injected 2 hours after the admission of the patient who remained fasting. Maximal urine concentrating ability was defined as the highest value of urine osmolality among the 8 measures performed on urine collected every 30 minutes.

### Kidney MRI

We performed a standardized protocol using T2 weighted sequence single-shot fast spin-echo (SSFSE). T2 SSFSE is an ultrafast MRI technique less susceptible to kinetic or respiratory artifacts and highly T2 weighted sequence allowing an optimal contrast between microcysts and renal parenchyma.

Kidney length was measured pole-to-pole. Cortex thickness was measured in the sagittal plane over the medullary pyramid facing the superior calyx, perpendicular to the cortical surface. Renal microcysts were defined as small (1–2 mm) round cystic lesions and quantified in both kidneys using a semi-quantitative scoring as previously described (8). MRI voxel values were not normalized using the cerebrospinal liquid as all patients were examined on the same machine GE 3T and as voxel value-based features were excluded from the analysis.

Manual segmentation of renal parenchyma excluding sinus, was performed using ITK SNAP 3.8.0. The software package has an interactive viewer that allows visualization in coronal, sagittal and axial planes (Figure 2). Finally, we extracted radiomic features using the pyradiomic module on 3D Slicer software 4.13.0. allowing the extraction of 107 first and second order features (17,18).

**Figure 2:**
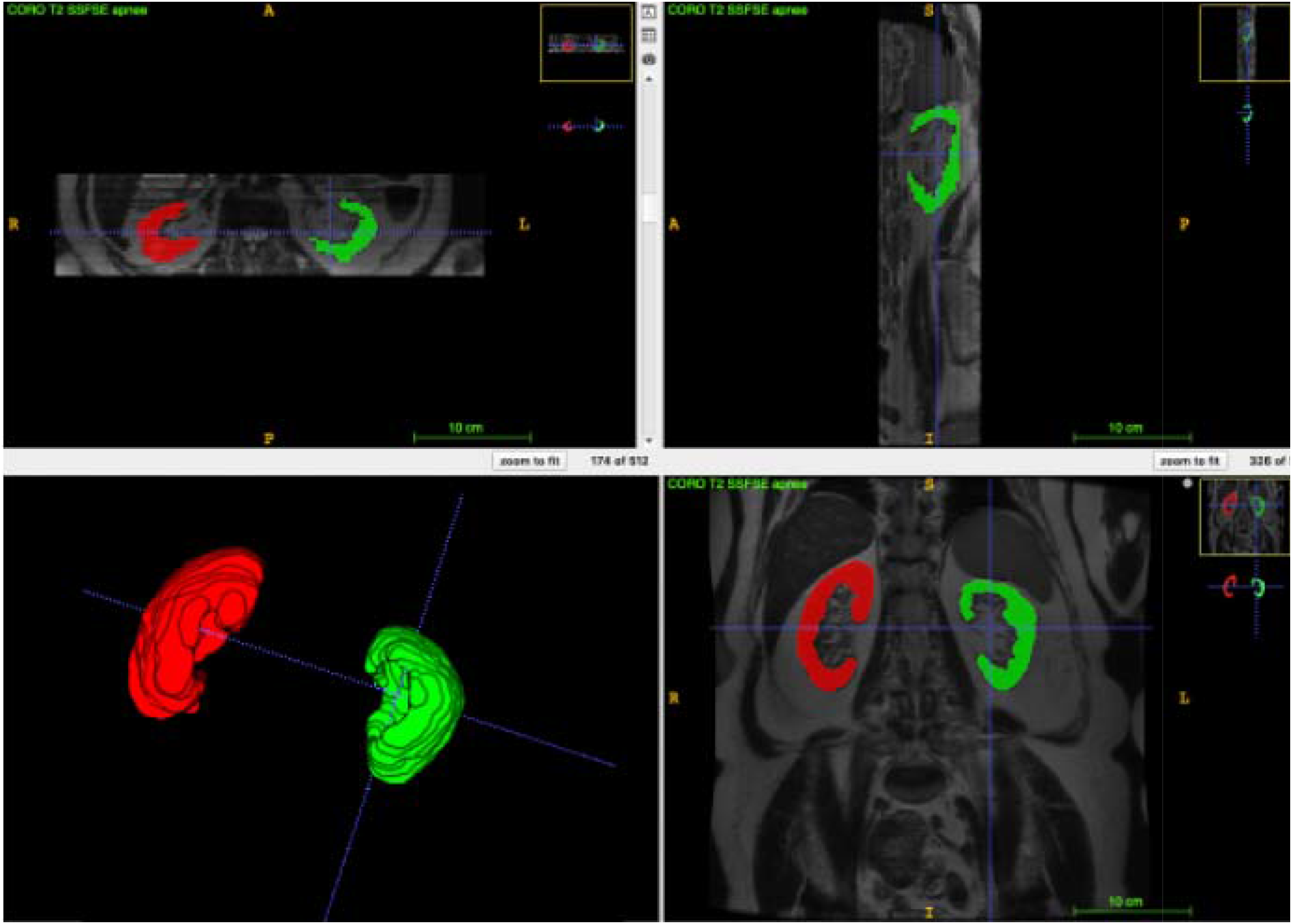
MRI manual renal segmentation on ITK SNAP 3.8.0.

### Statistical analyses

Using a randomization program, the cohort was randomly split into a training cohort (n=54) and a validation cohort (n=55). All radiomic features were analyzed and selected using penalized regression Lasso method (19). A Receiver operating characteristic (ROC) Curve was plotted to measure the Area Under the Curve (AUC) and the Youden Index allowed identifying the most accurate cut-off value for the detection of an mGFR below 60 ml/min/1.73m^2^. Two first-order features relevant for the prediction of an mGFR below 60 ml/min/1.73m^2^ were identified, namely skewness and kurtosis, and the second-order features of the grey level co-occurrence matrix (GLCM) that were identified were correlation and Cluster-Shade. Based on these features we built a quantitative score using fitting generalized linear model, used as a texture analysis index.

Categorical variables were shown as frequencies and percentages, quantitative variables were described as median (quartile 1-quartile 3). Patients’ characteristics were shown according to the textile analysis index above or below the median, and compared using the Chi-square test for categorical variables and the Mann-Whitney test for quantitative variables.

Determinants of texture analysis index were assessed using a multivariable linear regression stepwise model. Models were compared by maximum likelihood ratio test (nested models). Validity conditions of the multiple linear regression model were checked. First, the model’s suitability was tested using the Rainbow test. We then studied the residuals. Their independence was checked using the Durbin-Watson test associated with a graphical method. A quantile-quantile plot was made to check their normal distribution. The distribution’s homogeneity has been checked graphically by representating the square root of standardized residuals versus fitted values. The following covariates were included *a priori* in the model: age, gender, hypertension, lithium treatment duration in years, mGFR, serum vasopressin and the interaction term between lithium treatment duration and mGFR.

A two-sided p value < 0.05 was considered statistically significant. Statistical analyses and graphs were performed using GraphPad Prism 9.0 and R 3.4 software.

## Results

### Texture analysis index internal validation and accuracy to detect mGFR < 60 ml/min/1.73m^2^

We built a texture analysis index based on the first and second-order features described above. This quantitative index was tested using an AUC to detect an mGFR < 60 ml/min/1.73m^2^ (figure 3). The AUC was 0.85 in the training cohort and 0.71 in the validation cohort, with a Youden Index at 0.66 in the training cohort and 0.67 in the validation cohort, showing a satisfying internal validation of the technique.

**Figure 3:**
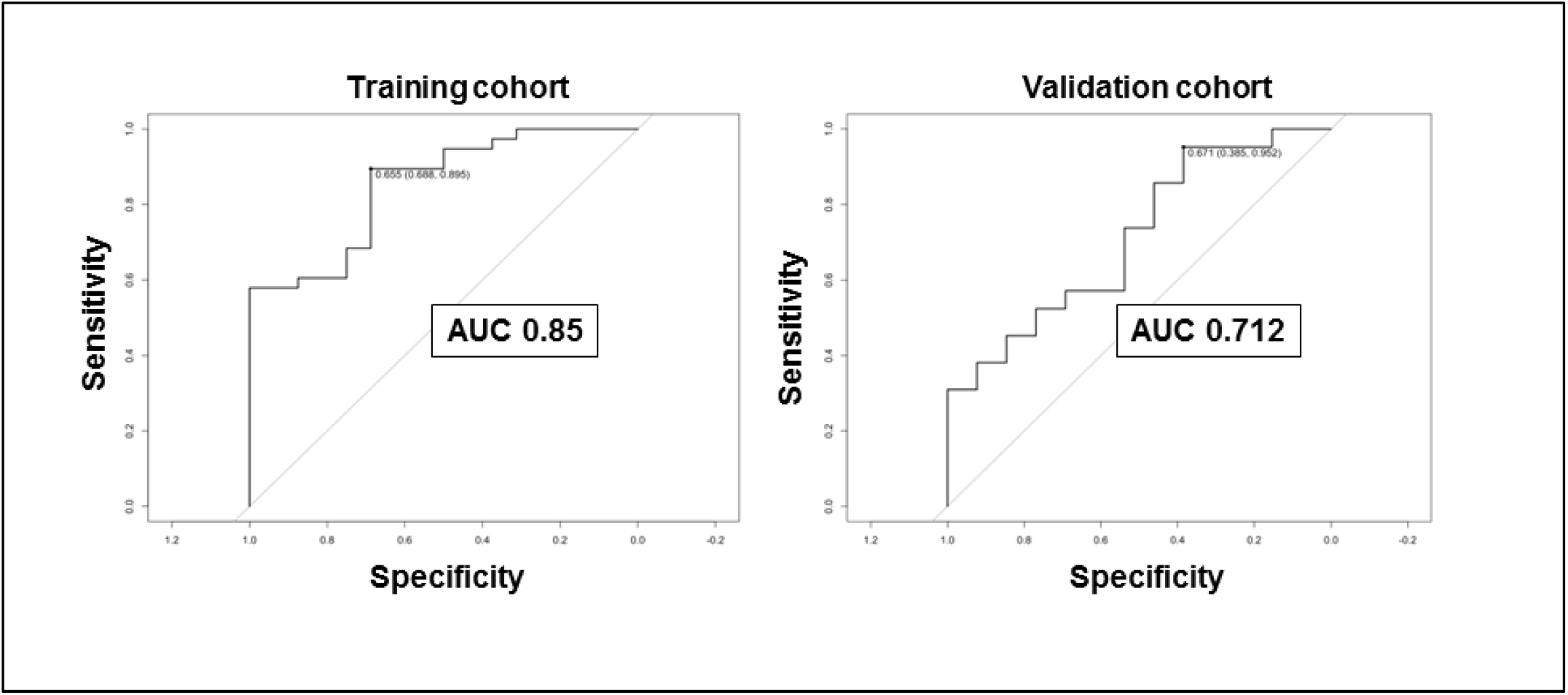
Receiver operating characteristic (ROC) Curve to detect mGFR < 60 ml/min/1.73m^2^ in the training and the validation cohort.

### Characteristics of the population

Characteristics of the studied population are reported in table 1. Median age was 51 [38-63] years in the entire cohort, median lithium treatment duration was 7 [2.6-16] years and median mGFR was 77.7 [56.2-90.3] ml/min/1.73m^2^. Compared to patients with a texture analysis index above the median, patients with a texture analysis index below the median were older (46 [34-54] versus 59 [42-66] years, p=0.001), with a higher body mass index (BMI) (25.5 [22.6-27.4] versus 27 [24.3-29.6] kg/m^2^, p=0.01), had a longer lithium treatment duration (6 [2-10.3] versus 10 [3.4-22.3] years, p=0.02), a lower daily lithium dose (800 [720-1200] versus 750 [500-1000] mg/d, p=0.04), a lower maximal urine concentrating ability (701 [573-856] versus 584 [328-793] mOsm/kgH_2_O, p=0.01) and more renal microcysts.

**Table 1:** Population characteristics in higher and lower structure index groups.

|  | overall<br>population | ≥median structure<br>index | <median structure<br>index | p-value |
| --- | --- | --- | --- | --- |
|  | n= 108 | n=54 | n=54 |  |
| Age, years | 51 [38-63] | 46 [34-54] | 59 [42-66] | **0.001** |
| Male | 43 (40) | 20 (37) | 23 (43) | 0.6 |
| BMI, kg/m<sup>2</sup> | 26 [23-29] | 26 [23-27] | 27 [24-30] | **0.01** |
| Hypertension | 29 (27) | 13 (24) | 16 (30) | 0.5 |
| Diabetes | 3 (3) | 1 (2) | 2 (4) | - |
| Hypothyroidism | 33 (31) | 15 (28) | 18 (33) | 0.5 |
| Lithium treatment duration, years | 7 [2.6-16] | 6 [2-10.3] | 10 [3.4-22.3] | **0.02** |
| Sustained-release formulation | 68 (63) | 34 (63) | 34 (63) | 1 |
| Lithium dose, mg/day | 800 [600-1000] | 800 [720-1200] | 750 [500-1000] | **0.04** |
| Serum lithium level, mmol/l | 0.7 [0.6-0.9] | 0.7 [0.5-0.8] | 0.7 [0.6-0.9] | 0.3 |
| mGFR, ml/min/1.73m<sup>2</sup> | 78 [56-90] | 83 [71-94] | 66 [46-84] | **<0.001** |
| ACR, mg/mmol | 1.4 [0.9-2.6] | 1.4 [0.9-2.3] | 1.5 [0.9-2.6] | 0.7 |
| Urine output, l/day | 2.3 [1.7-3.2] | 2.4 [1.9-3.2] | 2.3 [1.6-3.5] | 0.5 |
| Maximal urinary osmolality, mOsm/kgH<sub>2</sub>O | 665 [480-839] | 701 [573-856] | 584 [328-793] | **0.01** |
| Serum vasopressin, pg/ml | 6.6 [4.4-12.3] | 5.7 [4.0-10.0] | 7.9 [4.6-13.4] | 0.08 |
| Kidney MRI |  |  |  |  |
| Number of cysts | 0 [0-0] | 0 [0-0] | 0 [0-2] | - |
| Number of microcysts |  |  |  | **0.002** |
| 0 | 53 (49) | 36 (67) | 17 (31) |  |
| 1-5 | 25 (23) | 9 (17) | 16 (30) |  |
| 6-10 | 6 (6) | 2 (4) | 4 (7) |  |
| 11-20 | 6 (6) | 3 (6) | 3 (6) |  |
| 21-50 | 6 (6) | 5 (9) | 4 (7) |  |
| 51-100 | 3 (3) | 1 (2) | 2 (4) |  |
| >100 | 9 (8) | 1 (2) | 8 (15) |  |
| Structure analysis index | 0.79 [0.66-0.86] | 0.86 [0.82-0.90] | 0.66 [0.48-0.74] | **<0.001** |

Categorical and continuous data are expressed in n (%) and in median [quartile 1- quartile 3], respectively. Patients' characteristics were compared across texture analysis index (above or below the median) using the chi-2 test for categorical variables and Mann-Whitney for quantitative variables. mGFR: measured glomerular filtration rate, ACR: urinary albumin to creatinine ratio, MRI: magnetic resonance imaging.

### Texture analysis index according to patients’ characteristics

Texture analysis index was correlated with mGFR and age. The association remained significant between texture analysis index and age in the absence of microcysts (r^2^=0.13, p=0.008) but not for mGFR (figure 4A-C). Regarding the relationship between texture analysis index and lithium treatment duration, the lowest tertile of texture analysis index was observed in the patients with the longest treatment duration (figure 4D). Interestingly, the proportion of texture index within the medium tertile was the highest in patients treated with lithium for less than 5 years compared to longer treatment durations, whereas index within the highest tertile was found in patients treated with lithium for 5 to 15 years.

**Figure 4:**
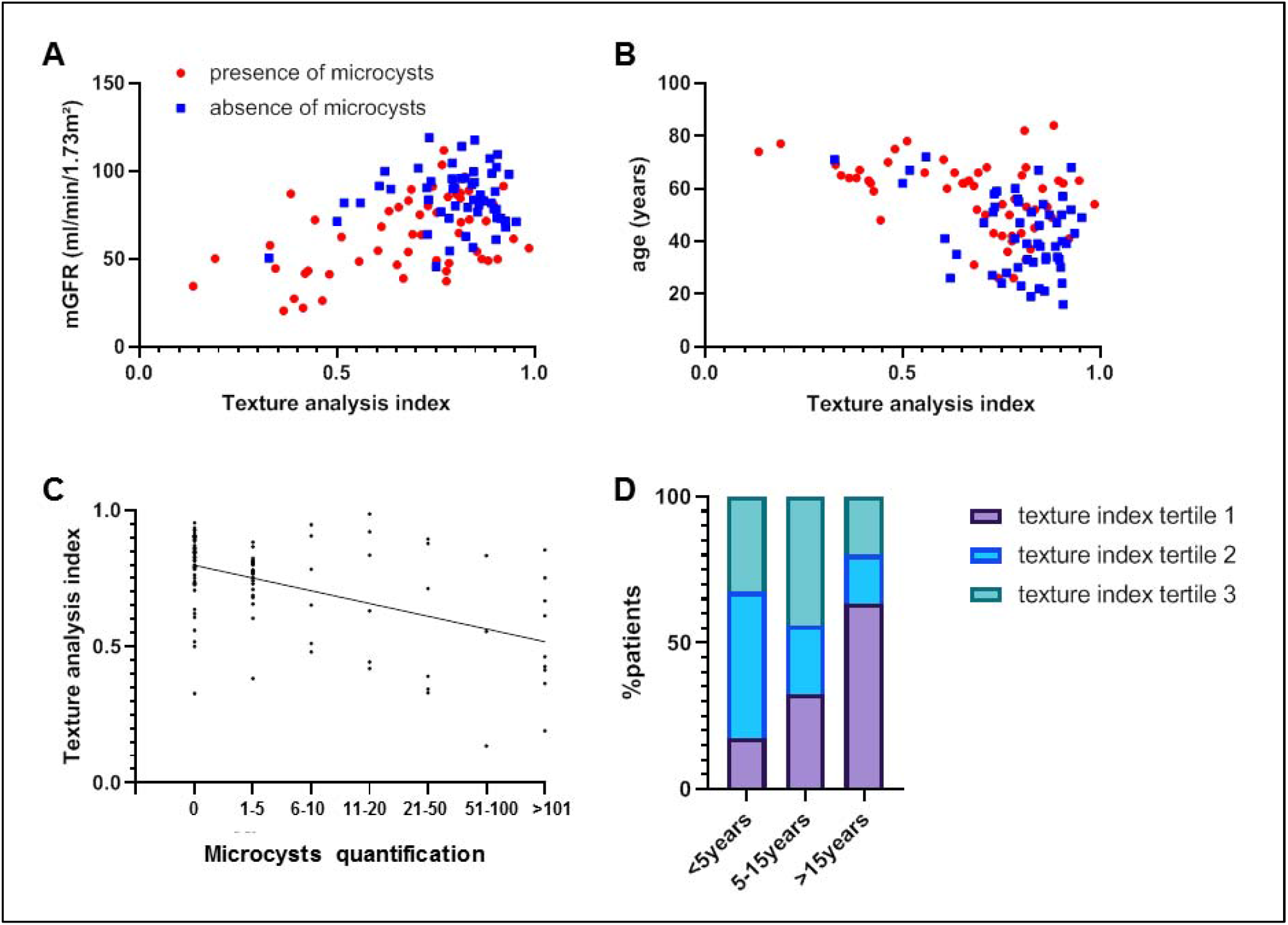
Correlations of texture analysis indexes with patients characteristics. Texture analysis index according to mGFR (A), to age (B) in patients with no microcysts (blue squares) and in patients with microcysts (red dots). Microcysts quantification according to texture analysis index (C). Texture analysis index tertiles according to lithium treatment duration (D).

### Determinants of texture analysis index

Texture index was negatively associated with age (β -0.004 ±0.001, p<0.001), lithium treatment duration (−0.01 ± 0.003, p=0.001) and serum vasopressin (−0.005 ± 0.002, p=0.02) independently of gender, albuminuria and hypertension (table 2). The association with mGFR vanished when lithium treatment duration was added to the model, but with a significant interaction between treatment duration and mGFR (p=0.01).

**Table 2.** Multivariable analysis of the determinants of texture analysis index.

| Dependent variable: texture index | $\beta \pm SE$ | p value |
| --- | --- | --- |
| **age** | -0.004 ± 0.001 | **<0.001** |
| **Lithium treatment duration** | -0.01 ± 0.003 | **0.001** |
| **mGFR** | -0.0008 ± 0.003 | 0.5 |
| **Serum vasopressin** | -0.005 ± 0.002 | **0.02** |
| **mGFR\*treatment duration interaction** | - | **0.01** |

Determinants were assessed using a multivariable linear regression stepwise model. The following variables were included in the model: age, gender, mGFR, lithium treatment duration, urine albumin/creatinine ratio, hypertension, interaction term between lithium treatment duration and mGFR.

## Discussion

This current study presents the first report of kidney MRI radiomics in patients treated with lithium carbonate. We built a texture analysis index that was associated with a decrease in mGFR. This texture index depended on lithium treatment duration, and was independently associated with age, vasopressin level and mGFR.

Lithium nephrotoxicity remains a major clinical issue because of its slow, asymptomatic and unpredictable evolution, which can lead to end-stage kidney disease (20). The benefit of lithium discontinuation and in particular the reversibility of renal function alteration is currently debated, as well as the probable individual susceptibility (9,20,21). Improved markers are therefore needed in these patients.

In our study, texture features were extracted from MR images and evaluated for their individual performance in detecting a mGFR below 60 ml/min/1.73m^2^. The first step was to validate the most relevant variables by means of an internal validation.

In a second step, we established a quantitative index allowing us to put it into the perspective of the patients’ characteristics. We observed an association between this index and the duration of lithium exposure. The number of microcysts was also correlated with the texture analysis.

Multivariable analysis of the determinants of this index showed a negative association between texture index and age, independently of lithium treatment duration. One could hypothesize that ageing might modulate kidney texture, raising the potential interest of this marker when studying structural changes associated with renal senescence.

Texture analysis index was also negatively associated with serum vasopressin in our population. Previous research reported the potential adverse effects of vasopressin on CKD progression in various experimental models (22), as well as in polycystic kidney disease (23). Elevated vasopressin levels in patients treated with lithium salts might reflect renal resistance to the action of vasopressin, eventually leading to decrease urine concentrating ability and nephrogenic diabetes insipidus (4). In line with the pathophysiology of polycystic kidney disease in which vasopressin-dependent V2R signaling induces cellular proliferation and cyst development (24), these findings raise interesting hypotheses regarding the pathophysiology of lithium-associated microcystic tubulo-interstitial nephropathy. Interestingly, previous report from Kline *et al*. showed that image texture change during PKD predicted CKD progression (25).

Finally, the texture index was negatively associated with lithium treatment duration. Interestingly, the association with mGFR in multivariable analysis disappeared when lithium treatment duration was added to the model. Moreover, a significant interaction was found between lithium treatment duration and mGFR, suggesting that mGFR influences the relationship between kidney texture and lithium exposure. This result should be put in light with our previous report showing an independent and strong relationship between mGFR and lithium treatment duration (8). However, the causative role of each variable is not yet demonstrated. Besides the work from Kline *et al*. on PKD (25), recent literature has investigated kidney texture analysis in kidney diseases such as glomerulonephritis (26) or radiation-induced kidney damage (27), as well as during CKD (28,29). Berchtold *et al*. (30) and Friedli *et al*. (31) have developed an MR imaging tool to detect renal fibrosis using an apparent diffusion coefficient, demonstrating the need for non-invasive diagnostic tools in CKD. To the best of our knowledge, there is no report of the association of histologically proven kidney interstitial fibrosis and radiomics analysis. Further research is thus needed to investigate this association globally during CKD.

We can identify some limitations to our study. Firstly, our study included a small sample of patients, which nevertheless remains the largest cohort of lithium treated patients with a standardized MRI protocol. Our study also lacks external validation on a control population due to the specific MRI protocol used in these patients. Finally, the cross-sectional design of our study does not allow evaluating the prognostic impact of our new biomarker. Our methodological validation with a gold-standard method of GFR measurement might thus be useful to future investigations including longitudinal follow-up of patients treated with lithium as well as patients with CKD.

In conclusion, we developed and validated a non-invasive renal texture-based imaging index that may be relevant in the diagnosis and prognosis of lithium-induced renal toxicity, as well as ageing and chronic kidney disease. It might also provide insights on pathophysiological mechanisms leading to lithium nephrotoxicity in future studies.

## Data Availability

All data produced in the present study are available upon reasonable request to the authors.

## Disclosures

The authors declare no conflict of interest.

## Funding

None.

## Acknowledgements

The authors wish to acknowledge the patients who accepted to enter the study, the nursing and medical staff who took care of the patients, and the technical staff who performed the imaging and the radioisotope measurements.

## Contributions

PB, MB, AD and NT designed the study. PB wrote the first draft of the manuscript. PB, LM, PF, EM, FB, BE, EVP, FV, MF and NT collected data. MB, LC and NT analyzed the data. PB conceived figure 1 and 2. MB conceived figure 3. NT conceived figure 4. All the authors contributed to the analyses of the data and edited the manuscript.

## Figure and Table Legends

**Supplemental table.** MR Imaging parameters.

|  |  |
| --- | --- |
|  | T2 SSFSE |
| Plane | Coronal |
| Repetition time (ms) | 1700-1850 |
| Echo time (ms) | 85-95 |
| Flip angle (°) | 90 |
| Fat suppression | No |
| Slice thickness (mm) | 3 |
| Intersection gap (mm) | 0 |
| Number of slices | 27-33 |
| Matrix size | 512 x 320 |
| Number of signal averages | 1 |
| Acquisition time (sec) | 145- 160 |

T2 SSFSE indicates breath-hold T2-weighted sequence single-shot fast spin-echo.

## Notes

### Competing Interest Statement

The authors have declared no competing interest.

### Funding Statement

This study did not receive any funding

### Author Declarations

IRB of APHP. Nord gave ethical approval for this work (approval #CER-2021-74)

